# Rural Medical Centers Struggle to Produce Well-Calibrated Clinical Prediction Models: Data Augmentation Can Help

**DOI:** 10.1101/2025.06.16.25329699

**Authors:** Katherine E. Brown, Bradley A. Malin, Sharon E. Davis

**Affiliations:** Vanderbilt University Medical Center, Nashville, TN

## Abstract

Machine learning models support many clinical tasks; however, challenges arise with the transportability of these models across a network of healthcare sites. While there are guidelines for updating models to account for local context, we hypothesize that not all healthcare organizations, especially those in smaller and rural communities, have the necessary patient volumes to facilitate local fine tuning to ensure models are reliable for their populations. To investigate these challenges, we conducted an experiment using data from a real network of hospitals to predict 30-day unplanned hospital readmission and a simulation study using data from a multi-site ICU dataset to evaluate the utility of synthetic data generation (SDG) to augment local data volumes. Several factors associated with rurality were correlated with model miscalibration and rural sites failed to meet sample size requirements for local recalibration. Our results indicate that deep learning approaches to SDG produced the best local classifiers.

## Introduction

Artificial intelligence (AI) tools, including machine learning models, have increased in their adoption for a variety of clinical tasks, including predicting the likelihood of disease^1^ or efficacy of medication^2^ as well as assessing the likelihood of hospital discharge^3^ or readmission^4^. Still, there are non-trivial challenges regarding the transportability of these models across sites^5–7^. A model trained at one site (i.e., hospital, floor, or ward) can deteriorate in overall reliability – measured as the calibration of model outputs to true likelihood of experiencing the outcome – when applied at other sites^8,9^. Various guidelines recommend updating machine learning models using data local to a site to account for this lack of transportability^10–12^; however, we hypothesize that not all healthcare organizations, especially those in smaller and rural communities, have the patient volume necessary to facilitate a meaningful update.

A significant portion of the US population resides in a rural area. As documented in the American Community Survey (ACS, 2018-2022)^13^, approximately 56,000,000 people (∼18% of the US population) reside in a ZIP Code Tabulation Area (ZCTA) designated as rural or borderline rural by the Centers for Medicare and Medicaid Services (CMS)^14^. Austen and colleagues^15^ propose a framework to evaluate clinical prediction models across multiple geographic sites, there has been little investigation into the impact of rurality on model performance or the capabilities of smaller medical centers to update clinical prediction models for their local populations. As such, there is an urgent need to characterize the impact and applicability of model updating and recalibration guidelines on the smaller medical centers that serve these patients. While Austen and colleagues^15^ propose a framework to evaluate clinical prediction models across multiple geographic sites, there has been little investigation into the impact of rurality on model performance or the capabilities of smaller medical centers to update clinical prediction models to fine-tune to their local populations.

Thus, in this work, we illustrate a proof-of-concept regarding the existing capabilities of rural medical centers to update clinical prediction models to ensure models are sufficiently well-calibrated for their patients. Additionally, in a simulation study, we evaluate the possibility of synthetic data generation^16,17^ to augment existing local data as an option to mitigate sample size concerns, extending previous studies utilizing data augmentation to mitigate equity concerns in AI models^18,19^. We specifically investigate the following research questions (RQs): **RQ 1:** What is the impact of rurality on model discrimination and calibration? **RQ 2:** How well can satellite healthcare facilities update clinical prediction models based on existing guidelines? **RQ 3:** How well can data augmentation mitigate sample size concerns when updating clinical models at smaller/rural sites compared to global data available across all sites?

## Methods

This study was approved by the Vanderbilt University Medical Center IRB (#240126).

Let us assume hospital *H*_0_ represents the main hospital or academic medical center associated with a network of hospitals, while hospitals *H*_1_, *H*_2_, …, *H*_*n*_ are satellite hospitals focused on the care of local communities (Figure 1). Let 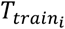 represent the set of patients at *H*_*i*_ during the model’s training period and 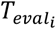 be the set of patients at *H*_*i*_ during the model’s evaluation period. Let *m* be the model trained on 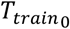.

**Figure 1.**
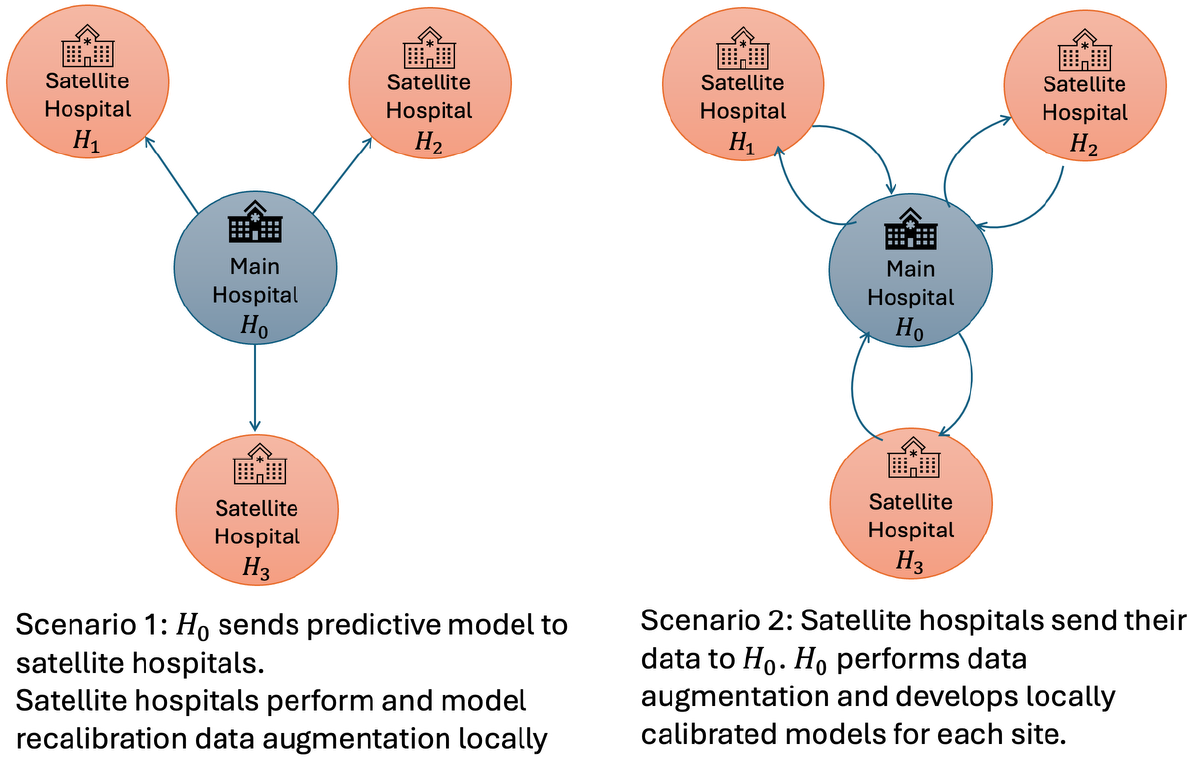
Schematic depicting the scenarios for synthetic data generation.

The model’s prediction on a patient *x*, denoted a *m*(*x*), can be further considered as the composition of *l*(*f*(*x*)). In this composition, *f* denotes a machine learning algorithm, such as a gradient-boosted tree or neural network, trained to produce a value corresponding to the logit of the likelihood of the event of interest. This logit is then provided to a calibrator model *l*(⋅) that produces the calibrated probability of the event of interest. Commonly, *l* is a logistic regression model. While the machine learning model *f* remains static after initial training, we can compose different calibrator models depending on the data used. For example, a calibrator derived from data from 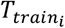 (e.g., *H*_*i*_) will be denoted as *l*_*i*_(⋅), and a calibrator derived from combining the training data from all sites (e.g., 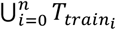) will be denoted as *l*_*A*_(⋅). Riley et al.^10,11^ derived several guidelines to determine the sample sizes needed to estimate recalibration parameters *l*_*i*_.

Given this framework, we can formally describe our main hypotheses as follows: **Hypothesis 1 (RQ 1)**: For sites *H*_1_, *H*_2_, …, *H*_*n*_, the optimal calibrator for a machine learning algorithm will not be *l*_0_(⋅) (e.g., the calibrator derived from the training data of the main hospital/site). **Hypothesis 2 (RQ 1)**: There is an association between rurality and model performance. **Hypothesis 3 (RQ 2)**: Hospitals *H*_1_, *H*_2_, …, *H*_*n*_ will not have the required amount of data in 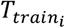 to produce the optimal calibrator *l*_*i*_(⋅). **Hypothesis 4 (RQ 3)**: Augmenting existing data in 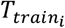 will produce a more optimal *l*_*i*_(⋅) compared to *l*_*A*_(⋅).

### Experiment 1: Evaluation of Real-World Clinical Prediction Model at Rural Sites

For the remainder of this study, we refer to this experiment as the Real-World Evaluation.

#### Data and Model

To characterize the ability of local sites to update and recalibrate clinical prediction models, we consider a model currently in production in a network of hospitals in the Middle Tennessee area (see Table 1). This network includes one urban academic medical center and two satellite hospitals in predominantly rural communities. Table 1 presents each site along with the population density of the county in which the site is located, area deprivation index^20^ (ADI), and whether or not the site is eligible for Health Resources and Services Administration (HRSA) rural health funding^21^. The model considered is the LACE+ Index^22^ which predicts unplanned 30-day readmission or mortality. We collected the highest LACE+ prediction during each index admission (i.e., not an unplanned readmission) for adult patients at each site between August 2021 and December 2024 and whether the patient experienced an unplanned readmission in the 30 days following discharge.

**Table 1.**
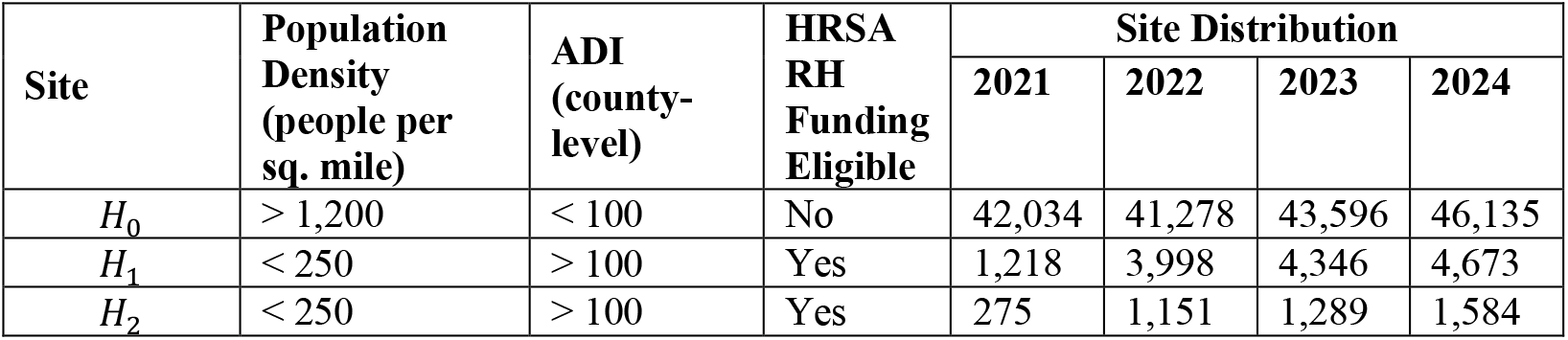
Distribution of patient cohorts, generalized area deprivation Index (ADI), population density to prevent identifiability of the sites. A higher ADI indicates higher disadvantage. Note: RH = Rural Health.

| Site | Population Density (people per sq. mile) | ADI (county-level) | HRSA RH Funding Eligible | Site Distribution |  |  |  |
| --- | --- | --- | --- | --- | --- | --- | --- |
|  |  |  |  | 2021 | 2022 | 2023 | 2024 |
| $H_0$ | > 1,200 | < 100 | No | 42,034 | 41,278 | 43,596 | 46,135 |
| $H_1$ | < 250 | > 100 | Yes | 1,218 | 3,998 | 4,346 | 4,673 |
| $H_2$ | < 250 | > 100 | Yes | 275 | 1,151 | 1,289 | 1,584 |

#### Experimental Methodology

Since we collected model predictions and outcomes from 2021 to 2024, we perform our evaluation temporally. For example, the experiment labeled “21-22” uses data from 2021 for calibration and data from 2022 for validation. Using the previously defined framework, we consider the model *l*_0_(*f*(*x*)) calibrated on data from *H*_0_. We also consider *l*_*A*_(*f*(*x*)) which is derived using aggregate data from all sites. Finally, we consider models calibrated on data from individual sites *l*_*i*_(*f*(*x*)). All calibrator models are implemented using Platt’s method^23^. We use Riley’s sample size guidelines^10^ for models with a binary outcome to determine if each individual site can recalibrate with existing data. If not, we resample the existing data at random to reach sample size requirements. Since our prediction task is binary, we use the C statistic to measure overall model discrimination. We calculate the calibration intercept *α* and slope *β* where ideal values are *α* = 0 and *β* = 1, respectively.

We also evaluate the correlation, if any, between common factors associated with rurality and model performance based on Pearson’s correlation coefficient. For the factors associated with rurality, we considered population density and ADI. Population density is often used as a defining characteristic for determining if a geographic area is rural^**24,25**^. ADI comprises of measures related to income, housing status, educational, employment status of residents in a geographic area^**20**^ and is frequently associated with health in the rural US^**26,27**^. In addition to measuring correlation with ***α, β***, and C, we also consider the magnitude of difference between ***α*** and ***β*** to their ideal values (i.e., |***α***| and |***β*** − **1**|) to determine if miscalibration is associated with rurality.

### Experiment 2: Simulation of Data Augmentation to Mitigate Sample Size Concerns

To characterize the efficacy of data augmentation to mitigate these sample size concerns, we developed a simulation using a publicly available critical care database that contains admissions across multiple sites nationwide^28^. We opted to use this external database since we did not have direct access to the all model inputs in the Real-World Evaluation. We assume a framework where a network of hospitals develops a machine learning model using data from the main site and then optimize the calibration of this model at the smaller community of hospitals within the network. In this experiment, we develop a model *l*_0_(*f*(*x*)) using data from the main site (*H*_0_). We then evaluate the impact of developing calibrator models *l*_*i*_ at each of the satellite hospitals *H*_*i*_ using data augmentation techniques in the form of oversampling and synthetic data generation^18,19^ to achieve necessary data volumes and compare these local calibrators to a global calibrator *l*_*A*_ using data from all sites. For the remainder of this study, we refer to this experiment as the Simulation Study.

#### Data

We perform our analysis with data from the eICU Collaborative Research Database^28^. This database contains de-identified ICU admissions from 2014 and 2015 for patients at 208 sites across the US. Each site is affiliated with an aggregate indication of its number of beds (e.g., > 500, 250-499, 100-249, < 100) and US Census region (i.e., South, Northeast, Midwest, West). We combine individual sites based on size (e.g., all hospitals in each region with more than 500 beds are considered as the main site or *H*_0_ for that region). For our simulation, we consider sites in the Northeast and South. In the southern US, there is a wide range in data volumes across hospital sizes; whereas, in the northeastern US, there is less variation in the number of patients by hospital size. Table 2 provides the size of each cohort based on hospital size and region. Patients with a discharge year of 2014 were partitioned into 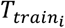 and patients with a discharge year of 2015 were partitioned into 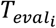. The eICU database contains features necessary to compute APACHE IV (Acute Physiology, Age, and Chronic Health Evaluation) hospital mortality prediction score. Using this features included in this score, the prediction task for our simulation is ICU mortality.

**Table 2.**
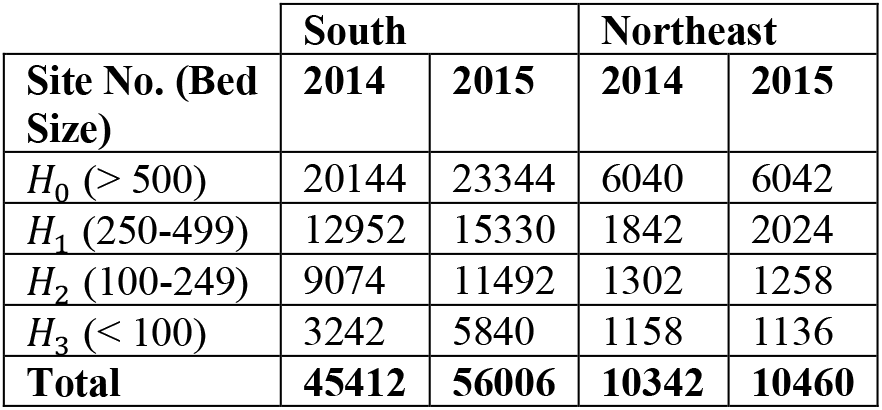
Distribution of patients per site based on US region.

#### Models

The classification model *f* in our simulation is a CatBoost gradient-boosted tree^29^ trained to predict ICU mortality based on the APACHE features. This model is trained using 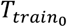 – patients from *H*_0_ with a discharge year of 2014 (see Figure 1). We use a random 75% of the data from *H*_0_ in 2014 to train the gradient-boosted tree to serve as our clinical prediction model to mitigate the likelihood of overfitting. The 25% unseen patients from *H*_0_ in 2014 are used to train the calibrator *l*_0_. For sites *H*_1_, *H*_2_, and *H*_3_, we use all data from 2014 to train the calibrator *l*_1_, *l*_2_, and *l*_3_, respectively. The calibrator model *l*_*i*_ is implemented using Platt’s method^23^. For *H*_1_, *H*_2_, and *H*_3_, local calibrator models are trained on 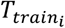 − patients from *H*_*i*_ with a discharge year of 2014 – with data augmentation if applicable. The aggregate calibrator model *l*_*A*_ is trained on all available data across sites from 2014.

#### Data Augmentation

Let *t*_*i*_ be the number of additional samples determined to be necessary to augment 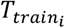 to produce a proper calibrator model at *H*_*i*_. In the event that a site does not have the requisite number of samples for proper calibration, we hypothesize that data augmentation in the form of oversampling or synthetic data generation may reduce this burden on smaller medical centers and facilitate updating to improve performance.

Modern synthetic data generation is dependent on modeling the univariate and multivariate distributions per feature (e.g., Gaussian Copula^30^) or by deep neural networks such as generative adversarial networks (GANs) and variational autoencoders (VAE)^17,31^. GANs commonly consist of two deep neural networks: a generator network that generates plausible synthetic instances and a discriminator network that determines if a data point is artificially generated. Alternatively, VAEs are built upon the autoencoders, which attempt to reconstruct data instances by reducing the representation to a latent space and expanding the latent representation into the reconstruction. Variational autoencoders build upon this network type by using a probabilistic model in the latent representation to encourage generation of synthetic instances. We choose to use a conditional GAN^32^ (CTGAN), Tabular VAE (TVAE)^32^, and Gaussian Copula (GaussianCopula)^30^ implemented in the Synthetic Data Vault for our analyses. The computational requirements for the proposed synthetic data generation may be difficult for smaller medical centers to realize. Thus, we consider four possible data augmentation scenarios with oversampling and synthetic data generation (Figure 1 and Table 3)^18,19^.

**Table 3.** Description of data augmentation methods in the Simulation Study. Note: SDG = Synthetic Data Generation.

|  | Name | Description |
| --- | --- | --- |
| Scenario 1: Local sites have computational resources supporting synthetic data generation | SDG | Train a synthetic data generator on $T_{train_i}$. Generate $t$ instances required to reach minimum sample size requirements. |
| | SDG + Oversample | Train a synthetic data generator on $T_{train_i}$. Generate $t/2$ instances from SDG. Oversample $T_{train_i}$ with replacement for the remaining $t/2$ instances. |
| Scenario 2: Local sites lack computational resources supporting synthetic data generation | Oversample | Oversample $T_{train_i}$ with replacement to reach the minimum sample size required. |
| | SDG, Conditional | Train a synthetic generator on $\bigcup_{i=0}^n T_{train_i}$. Sample $t$ total instances with the desired site. |

First, we consider the scenario when a satellite hospital *H*_*i*_ (with *i* ≥ 1) has access to the computational resources necessary to augment their data with synthetic instances (Figure 1, Scenario 1). In this scenario, the local site will train a synthetic data generation model on their original data. Then, the local site could sample the *t* additional examples needed from the synthetic data generator. As an alternative approach with oversampling, the local site could sample ⌈*t*/2⌉ observations from the synthetic data generator and oversample another ⌊*t*/2⌋ from 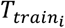 (sampling with replacement if necessary). Second, we consider the scenario when a satellite hospital *H*_*i*_ (with *i* ≥ 1) does not have access to the computational resources necessary to augment their data with synthetic instances (Figure 1, Scenario 2). Then, the local site would either be required to oversample from 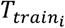 or be dependent on *H*_0_ to generate the necessary synthetic samples. In the latter approach, we assume *H*_0_ has access to data at all sites to train a common synthetic data generation model. Then, we sample from the generator to produce *t*_*i*_ samples from *H*_*i*_.

#### Experimental Methodology

We run the proposed simulation twice. Within each execution of the simulation, each data augmentation and respective recalibration is performed five times. This results in 10 measurements of calibration performance. Since our prediction task is binary, we use the C-statistic to measure overall model discrimination and measure calibration with the calibration intercept *α* and slope *β*, where ideal values are *α* = 0 and *β* = 1, respectively.

## Results

### Impact of Rurality on Model Performance

The Real-World Evaluation assessed the discrimination and calibration of the LACE+ Index at a network of hospitals in the Middle Tennessee area. These hospitals include one urban academic medical center and two rural satellite hospitals. Table 4 presents the C-statistic along with the coefficients ***α*** and ***β*** of the calibration line of the model developed using data from the main site ***H***_**0**_ and evaluated at all three sites ***H***_**0**_, ***H***_**1**_, and ***H***_**2**_. We compute the 95% confidence intervals based on the performance at three evaluation scenarios across a total of four years. We note that C-statistic – which indicates the model’s overall ability at identifying the outcome of interest – differs by as much as about 12% across the individual sites and ***H***_**0**_ does not have the best discrimination. Model calibration – which is measured by the slope ***β*** and intercept ***α*** of the calibration line formed by the model’s predictions – exhibits more noticeable differences across the sites. When using a calibrator derived from the main site (Table 4, ***l***_**0**_), we found that rural sites experienced worse calibration than the primary urban site.

**Table 4.** C-statistics and calibration coefficients ***α*** and ***β*** across each year of calibration/evaluation based on calibrator.

| | Site | C (95% CI) | $\alpha$ (95% CI) | $\beta$ (95% CI) |
| --- | --- | --- | --- | --- |
| $l_0$ | $H_0$ | 0.63 (0.63, 0.63) | 0.1 (0.1, 0.11) | 1.05 (1.05, 1.05) |
| | $H_1$ | 0.68 (0.68, 0.68) | 0.88 (0.88, 0.89) | 1.59 (1.59, 1.59) |
| | $H_2$ | 0.56 (0.56, 0.56) | -0.76 (-0.77, -0.76) | 0.66 (0.66, 0.66) |
| $l_A$ | $H_0$ | 0.63 (0.63, 0.63) | 0.09 (0.09, 0.09) | 1.03 (1.03, 1.03) |
| | $H_1$ | 0.68 (0.68, 0.68) | 0.87 (0.86, 0.87) | 1.57 (1.56, 1.57) |
| | $H_2$ | 0.56 (0.56, 0.56) | -0.76 (-0.77, -0.76) | 0.65 (0.65, 0.66) |
| $l_i$ | $H_0$ | 0.63 (0.63, 0.63) | 0.1 (0.1, 0.11) | 1.05 (1.05, 1.05) |
| | $H_1$ | 0.68 (0.68, 0.68) | 0.62 (0.6, 0.63) | 1.25 (1.24, 1.26) |
| | $H_2$ | 0.56 (0.56, 0.56) | 0.3 (0.28, 0.31) | 1.1 (1.09, 1.11) |
| $l_i$<br>$OS$ | $H_0$ | N/A | N/A | N/A |
| | $H_1$ | 0.68 (0.68, 0.68) | 0.21 (0.2, 0.22) | 1.04 (1.04, 1.05) |
| | $H_2$ | 0.56 (0.56, 0.56) | -0.3 (-0.33, -0.28) | 0.78 (0.77, 0.8) |

We found that model discrimination and model calibration curve coefficients themselves are not highly correlated with either factor of rurality (Table 5). For discrimination performance, this may be a by-product of the similarity of performance across included sites, and if other models or networks experience larger differences in discrimination across sites, this correlation may change. Interestingly, the magnitude of miscalibration (|***α***| and |***β*** − **1**|) was highly correlated with both considered factors of rurality. For population density, this is to be expected. Rural areas have lower population and as a result, lower population density. Since there are fewer people in the areas serviced by satellite medical centers, the overall patient population during a given period is expected to be smaller for satellite hospitals compared to their urban counterparts. A simplistic explanation is that fewer patients mean fewer samples that can be used for model calibration and validation, resulting in larger calibration error when a model is applied at satellite hospitals. Alternatively, population density may be a proxy variable for unmeasured or unmeasurable aspects of rurality. Additional, analyses will be necessary to confirm.

**Table 5.**
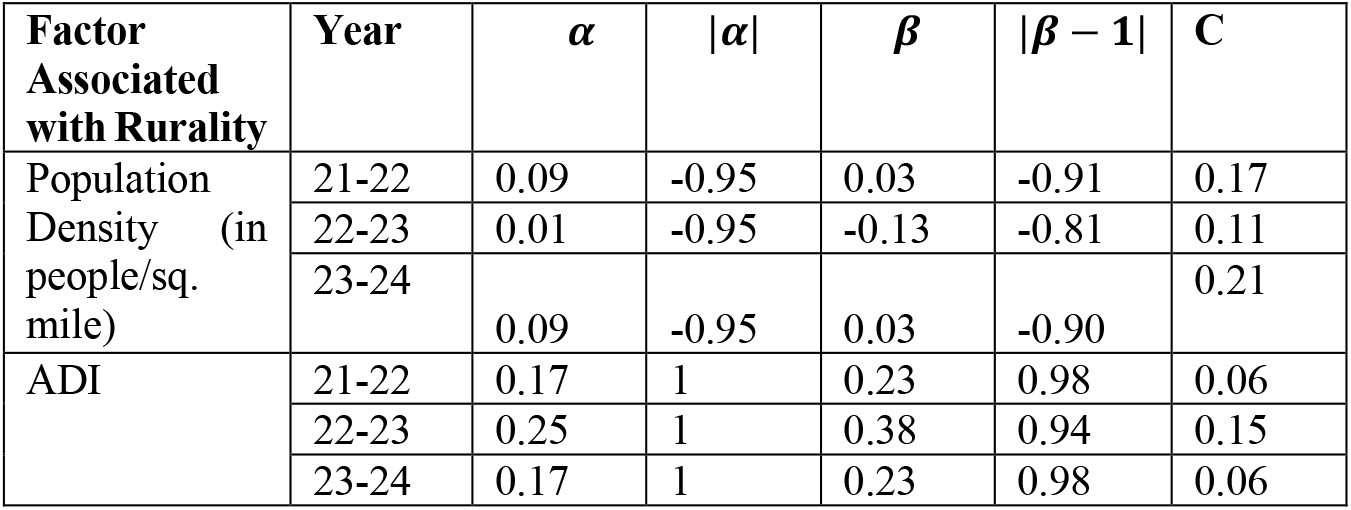
Pearson correlation coefficients for factors associated with rurality (population density and ADI) compared with model discrimination, calibration, and miscalibration.

The correlation with Area Deprivation Index is also expected; however, the explanation for this correlation is not as straightforward. We calculated the ADI for the county in which each site is located. Higher ADI (i.e., higher disadvantage) is commonly associated with predominantly rural areas. Since population density is often a defining characteristic of rurality, the correlation between model miscalibration and ADI could be a by-product of existing correlation between ADI and population density (***ρ*** = −**0. 97**). However, high ADI is not both necessary and sufficient to define a rural area (i.e., high ADI can occur in a predominantly urban area). Thus, another possible explanation is that sites located in areas with higher ADI (e.g., higher disadvantage) have the propensity to face greater model miscalibration concerns. Factors including accessibility of healthcare, potentially differing protocols or opportunities for follow-up, and different distributions of potential cases could result in models requiring more detailed maintenance than simple recalibration to best serve local populations. Further experiments with nationwide data across multiple models will be necessary to elucidate the nature of the relationship between ADI and model miscalibration.

#### Ability of Satellite Hospitals to Update Clinical Prediction Models

Additionally, in the Real-World Evaluation, we evaluated the capability of real-world data from a multi-hospital network to recalibrate a model at geographically distinct satellite sites. Table 6 provides the number of samples required for local site-specific recalibration, samples available, and the number of samples to be generated, if applicable. First, we consider using all available data at a local site, regardless of if the site has the requisite samples for local recalibration. We examined the C-statistic and calibration coefficient (***α*** and ***β***) values of the site-specific calibrators ***l***_**1**_ and ***l***_**2**_ (Table 4, ***l***_***i***_). We see mild improvement in calibration at ***H***_**1**_ when using available samples and more noticeable improvement in calibration at ***H***_**2**_ when using available samples. Of note, the range of possible values in the 95% confidence interval increases at both sites from 0.00-0.01 to 0.02 to 0.03 indicating that while better calibration occurred at these sites, the variation in calibration performance also increased. This is likely a result of neither ***H***_**1**_ or ***H***_**2**_ having the recommended number of samples for optimal, local recalibration^**10**^.

**Table 6.**
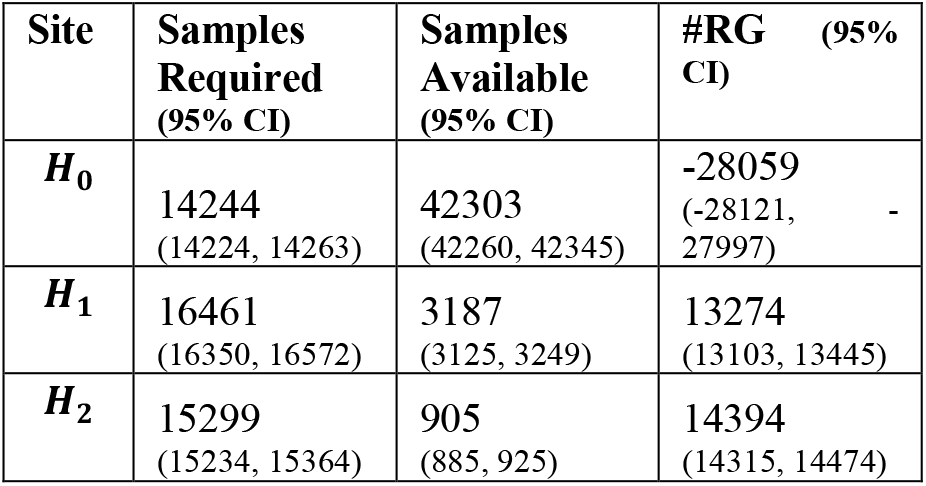
Sample size requirements for each site in the Real-World Evaluation, aggregated across time. Number of samples required to generate is denoted #RG.

Thus, we consider a scenario in which available data across all sites are aggregated to form a single, multi-site calibrator (Table 4, ***l***_***A***_). This approach resulted in calibration very closely aligned to using the calibrator derived from the main site, ***l***_**0**_. This is likely due to the fact that samples from site ***H***_**0**_ composes between 88% and 97% of available samples for recalibration for each year in which the data was available. This results in rural sites ***H***_**1**_ and ***H***_**2**_ having a low prevalence among samples used for recalibration. Ultimately, this indicates that global recalibration may be ill-suited when patients at rural sites have low prevalence in the overall sample space for recalibration.

Finally, we considered developing site-specific calibrators, ***l***_**1**_ and ***l***_**2**_, by oversampling (OS) available data to reach minimum sample size requirements (Table 4, ***l***_***i***_ ***OS***). We find that for site ***H***_**1**_, oversampling to reach sample size requirements was sufficient to successfully calibrate the local calibrator ***l***_**1**_. For site ***H***_**2**_, oversampling available data does not result in the same improvement in calibration as seen by ***H***_**1**_. We suspect this is a result of the fact that ***H***_**2**_ consists of less than 3% of all samples – the smallest site in our evaluated cohort. Thus, it is reasonable that the vast amount of oversampling required to reach minimum sample size requirements ultimately caused the calibrator to overfit.

#### Data Augmentation to Mitigate Sample Size Concerns

As evidenced in our findings from the Real-World Evaluation, it can be seen that satellite hospitals within a network may experience poor calibration when applying models calibrated on data from a primary hospital and oversampling to reach minimum sample size requirements may be insufficient for recalibration. To better emulate a local network of hospitals, we combined individual sites based on size (see Table 2). Additionally, we note in Table 2 that the northeast US has fewer patients in this database compared to the southern US. One satellite site in the southern US and all three satellite sites in the northeast US required data augmentation to reach the minimum sample size for recalibration.

Table 7 presents the C-statistic and 95% confidence interval when ***H***_**0**_ trains and calibrates a clinical prediction model on 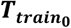 and provides this model to ***H***_**1**_, ***H***_**2**_, and ***H***_**3**_ without local recalibration. We note that there are minor differences in C-statistics across sites; however, we note moderate variability in calibration across different sites. Given the observed differences in calibration, we calculated the minimum sample size required for estimating recalibration parameters according to Riley and colleagues^10^ (Table 8). We note that for sites in the southern US, only ***H***_**3**_ requires data augmentation to meet sample size requirements, but for the northeast US, all three satellite sites ***H***_**1**_, ***H***_**2**_, and ***H***_**3**_ require data augmentation. We note that generating each of these sites required combining data from hospitals of similar sizes within each region into ***H***_**0**_, ***H***_**1**_, ***H***_**2**_, and ***H***_**3**_. Even in this artificial scenario, there are instances where recalibration cannot be adequately performed with the available sample sizes. This further motivates the need to understand the impact of data augmentation for recalibrating models at sites with limited sample sizes.

**Table 7.** C-statistic and 95% confidence interval of model on unseen data in the Simulation Study. Calibrator used is the main site calibrator, *l*_0_.

| Site No. | South |  |  | Northeast |  |  |
| --- | --- | --- | --- | --- | --- | --- |
| | C (95% CI) | $\alpha$ (95% CI) | $\beta$ (95% CI) | C (95% CI) | $\alpha$ (95% CI) | $\beta$ (95% CI) |
| $\mathbf{H}_0$ | 0.85<br>(0.85, 0.85) | -0.38<br>(-0.48, -0.28) | 0.79<br>(0.76, 0.81) | 0.81<br>(0.81, 0.82) | -0.57<br>(-0.58, -0.56) | 0.57<br>(0.57, 0.58) |
| $\mathbf{H}_1$ | 0.85<br>(0.85, 0.85) | -0.45<br>(-0.53, -0.37) | 0.79<br>(0.77, 0.81) | 0.81<br>(0.81, 0.82) | -0.84<br>(-0.85, -0.84) | 0.51<br>(0.5, 0.51) |
| $\mathbf{H}_2$ | 0.84<br>(0.84, 0.84) | -0.41<br>(-0.47, -0.35) | 0.85<br>(0.83, 0.86) | 0.81<br>(0.8, 0.82) | -1.09<br>(-1.18, -1) | 0.62<br>(0.6, 0.65) |
| $\mathbf{H}_3$ | 0.84<br>(0.84, 0.84) | -0.23<br>(-0.35, -0.11) | 0.84<br>(0.81, 0.87) | 0.79<br>(0.79, 0.8) | -0.87<br>(-0.92, -0.82) | 0.54<br>(0.53, 0.56) |

**Table 8.** Sample size requirements for each site in the Simulation Study, aggregated across time. Rows in bold indicate sites in which data augmentation is necessary. Number of samples required to generate is denoted #RG.

|  | South |  |  | Northeast |  |  |
| --- | --- | --- | --- | --- | --- | --- |
| Site | Samples Required | Samples Available | #RG | Samples Required | Samples Available | #RG |
| $H_0$ | 6007 | 8810 | N/A | 3395 | 2586 | 809 |
| $H_1$ | 6961 | 12952 | N/A | 3555 | 1302 | 2253 |
| $H_2$ | 6854 | 9074 | N/A | 4309 | 1842 | 2467 |
| $H_3$ | 5787 | 3242 | 2545 | 4645 | 1158 | 3487 |

Figure 2 presents a two-dimensional plot of the calibration intercept ***α*** on the x-axis and calibration slope ***β*** on the y-axis. The closeness of the point (***α, β***) is to (0,1) indicates the success of the recalibration.

**Figure 2.**
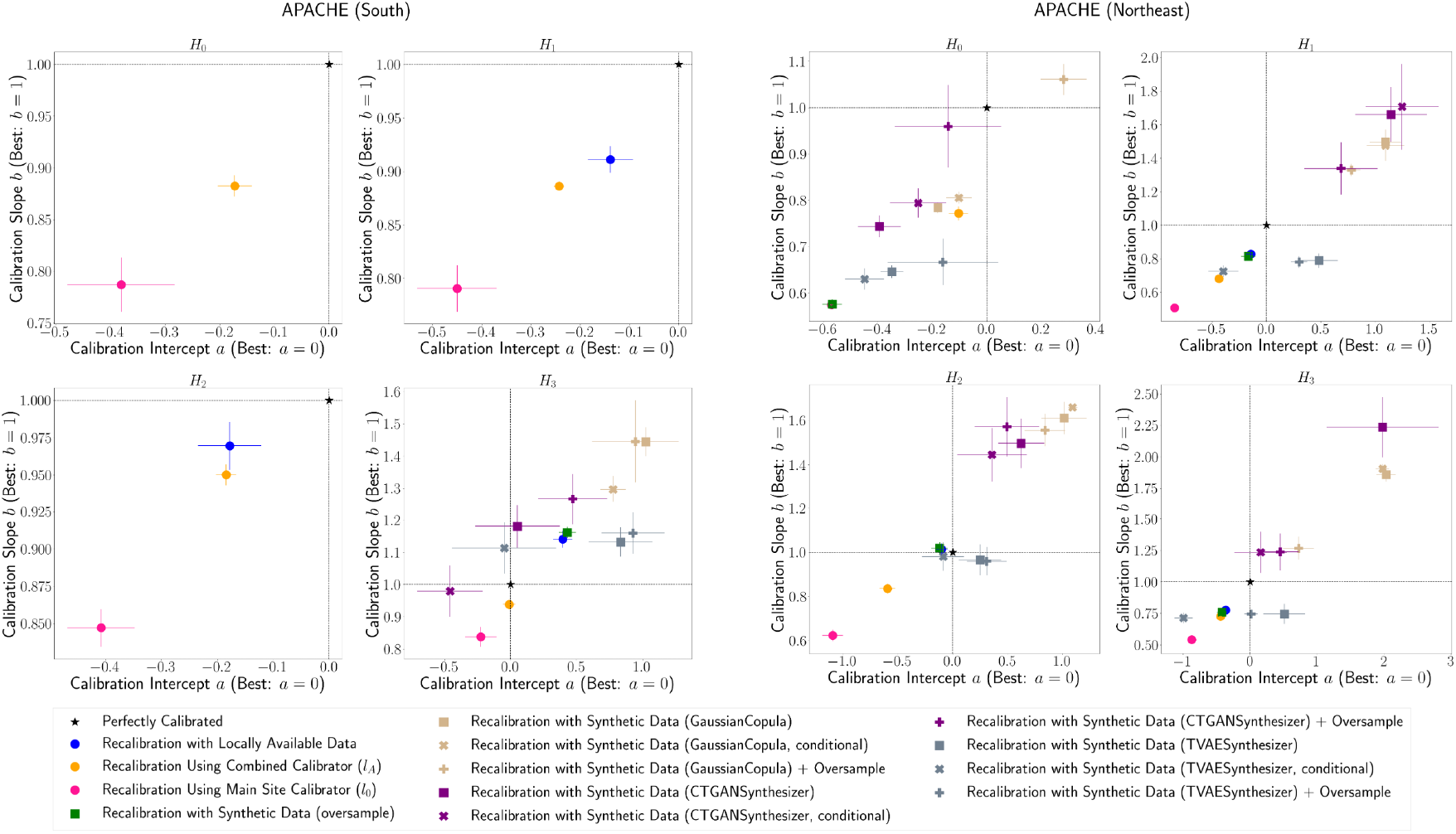
Plots of slope/intercept of the calibration curve for each data augmentation method.

For the APACHE model in the southern US, sites ***H***_**1**_ and ***H***_**2**_ had the requisite samples for recalibration, and we found that using locally available data for these sites yielded the best calibration on unseen data. Of note, ***H***_**2**_ performance displayed higher variance than ***H***_**1**_ and has a smaller number of available instances for recalibration and evaluation. This could be a result of an overall smaller pool of data used to evaluate calibration at this site. Only ***H***_**3**_ required data augmentation to reach sample size requirements, and we find that training CTGAN on data from all sites then conditionally sampling to retrieve instances for ***H***_**3**_ to yield the most calibrated predictions across data augmentation strategies.

For the APACHE model in the northeast US, we find that all three satellite sites, ***H***_**1**_, ***H***_**2**_, and ***H***_**3**_ required data augmentation to meet the sample size requirements for recalibration. For sites ***H***_**1**_ and ***H***_**2**_, oversampling or using locally available data resulted in the best performance, with conditionally sampled TVAE-generated data resulting in the best calibrator using synthetic data generation. For ***H***_**3**_, the combination approach using synthetically generated data and oversampled local data produced the best local calibrator. We note for this simulation scenario there is generally less available data than for the APACHE model in the northeastern US. This lack of data could result in the synthetic data generators not producing adequate data for recalibration and as a result recalibration efforts benefitting more from a combined approach of synthetic data and oversampling.

Both geographic regions had differing capabilities at the primary site ***H***_**0**_. In the southern US, the main site had the requisite samples for recalibration; however, calibration performance using the main site calibrator ***l***_**0**_ had higher variation than the combined calibrator ***l***_***A***_. For the northeast US, the main site did not have the requisite number of samples for local recalibration; however, we see that combining synthetically generated data and oversampling the existing data resulted in a better calibrated classifier than the combined calibrator ***l***_***A***_.

## Discussion and Conclusion

### Real-World Evaluation

Our analysis illustrates mild differences in discrimination and moderate deterioration in calibration when applying a model developed at a primary, urban site to satellite, rural sites. This is not unexpected in the context of previous studies^**8,9,15**^. Austin and colleagues^**15**^ developed validation techniques of prediction models across multiple sites. In their evaluation of a mortality prediction model for heart failure patients, they noticed more pronounced differences in C-statistic than in our study; however, we noticed more variation across sites in both calibration slope and intercept than their study. While this may be an artifact of the different patient cohort and prediction model examined in both studies, both studies highlight the growing need for consideration of local validation for models applied at multiple sites, even within the same network of hospitals.

While differences in the C-statistic were minimal for this model and scenario, further evaluation of other models at these sites and at other urban-rural hospital networks will be necessary to confirm the nature of intra-network performance variation. Conversely, we noticed more extensive calibration differences across geographic sites and suspect that this will be a trend for other models and networked hospitals. Still, additional external analysis is necessary to confirm. Nonetheless, we note that our findings are consistent with prior work that indicates calibration is more sensitive to differences between training and validation cohorts than discrimination^8,9^.

Lyons and colleagues^**33**^ conducted an external validation of a proprietary sepsis model across a network of nine hospitals. In their study, the evaluated model was developed by an EHR vendor and provided for immediate use by institutions. The model experienced pronounced differences in discrimination performance across sites. Additionally, the authors observed correlation between C-statistics and multiple variables associated with sepsis diagnosis, including time to diagnosis, sepsis incidence rates, comorbidity burden, and cancer prevalence. These factors, except time to diagnosis, were noted to be inversely correlated to model discrimination performance. Their findings and those from our study highlight the need to investigate clinical and socio-economic factors that could be associated with degradation in model discrimination and calibration.

### Simulation Study

In light of concerns posed when evaluating calibration of models across geographic sites, we performed a simulation study with a publicly available multi-site ICU database. We evaluated the efficacy of synthetic data generation using sites in the northeast and southern US. We find that for the southern US, deep learning-based approaches to generate sufficient data for local recalibration resulted in classifiers that were closest to a perfect calibration; however, for the northeast US, using existing data and oversampling was the most advantageous for two of the three satellite sites. Since the southern US had more data overall, it is reasonable to hypothesize that the deep learning approaches may have overfit the data in the northeast US, resulting in less robust synthetic instances. This hypothesis is further validated by Yan et al.^17^ who found that using deep learning approaches on samples and concepts with low prevalence may result in statistical properties being inadequately learned by GAN-based synthetic data generation techniques. Overall, though, despite concerns about smaller sample sizes used to train synthetic data generation techniques, deep learning-based approaches typically produced better samples for recalibration than the statistical synthetic data generation technique tested. This indicates that deep learning-based approaches hold more promise for producing samples for recalibration than statistical data generation techniques. Additionally, the benefits of using synthetic data generation to produce additional samples for recalibration is further demonstrated when the main site itself does not have the requisite samples for recalibration, as we observed for the northeast US (Figure 2).

Additionally, we find that recalibration was more successful using data generated by deep learning-based approaches when such generator models were trained on all available data as opposed to at individual sites (e.g., conditional sampling approach). While this would logically follow from our previous hypothesis and be the most accessible for sites with more limited computational or technical staffing resources, it poses potential concerns regarding data access and privacy. This conditional approach requires that the main site has access to data from each satellite site, which may not be true or feasible for some networks of institutions. This could lead to a situation where a site does not have the computational resources to perform their own synthetic data generation, but the main site does not have the necessary access privilege to perform the synthetic data generation. Thus, further research is necessary to understand when these scenarios occur and how to make synthetic data generation and other data augmentation methods more applicable for these sites.

Finally, guidelines for minimum sample size for estimating recalibration parameters also apply to determining the number of samples necessary to perform validation of model discrimination and calibration. Since the southern US had a higher data volume than the northeast US, the southern sites had the minimum sample size for recalibration and validation. The northeast US, however, did not meet the sample size requirements for robust validation (see Tables 4 and 9). Thus, another possible reason as to why synthetic data generation was not the best data augmentation technique could be due to an insufficient data volume for validation to provide a complete picture of model calibration and discrimination. Performing data augmentation for validation was outside of the scope of this work and require additional considerations to prevent data leakage and evaluate whether generated data are robust for model validation.

### Limitations and Conclusions

There are several avenues for future work to enhance the robustness of our findings. First, we only consider the scenario in which a machine learning model is trained using main site data. Future work should also consider the scenario with a model trained across all sites and other validation techniques for multi-site models^15^. Additionally, we focused on synthetic data generation with little fine-tuning, nor do we use a synthetic data generation method developed specifically for the purposes of synthetic EHR record generation. Finally, future work should consider larger cohorts like the All of Us Research Program^34^ and data from other multi-center hospital networks.

In this work, we explore the transportability of clinical prediction models across geographically diverse sites and the ability of small, rural healthcare centers to locally recalibrate models given available data resources. While clinical prediction tools can help realize increased health outcomes, the urban-rural divide in healthcare in the US also extends to the localization of clinical prediction models to smaller healthcare facilities. Through our analysis, we see that connections to large medical centers is not enough to promote accurate prediction models at all sites within a healthcare system, and data augmentation and synthetic data augmentation may provide the necessary data volumes to enable local recalibration at smaller facilities. Data augmentation approaches, however, may assume unrestricted access to local data volumes or local access to computational resources which may be infeasible for some healthcare networks. Addressing these needs will require novel approaches that balance local and network-supported data augmentation.

## Data Availability

The datasets generated and/or analyzed during the current study are not publicly available due to patient private information investigated but are available from the corresponding authors on reasonable request.

## Acknowledgements

This research was funded by the National Institutes of Health grants T15LM007450 and U54HG012510.

